# Social and Health Seeking Determinants of Antibiotic Use in Vietnamese Children Under 5: Analysis of National Household Survey Data

**DOI:** 10.1101/19013292

**Authors:** Tianyi Wang, Thanh Le Viet, Tung Trinh Son, Rogier van Doorn, Charlotte Zheng, Gerald Jamberi Makuka, Sonia Lewycka

## Abstract

**Background:** Antibiotic resistance is an important global public health issue, perpetuated by increases in antibiotic use. In low- and middle-income countries (LMICs), tackling antibiotic resistance bacteria is especially challenging. Due to high rates of infectious disease and continuing high mortality from untreated bacterial infections, policy must balance tackling both antibiotic access and antibiotic overuse. This paper investigates the social and health-seeking determinants that impact appropriate and inappropriate antibiotic use in Vietnamese children under 5 for Acute Respiratory Illness (ARI).

**Methods:** Descriptive analyses and logistic regression models were performed on country-wide household data from UNICEF Multiple Indicator Cluster Surveys in 2006, 2011, 2014.

**Results:** Results show that antibiotic overuse is higher in those who sought care from a healthcare provider than those who self-treated. In 2014, children who sought care at private facilities and government facilities were more likely to overuse antibiotics for mild respiratory infections (OR 6.1 and OR 3.8 respectively) than those who did not seek care at private and government facilities respectively. Furthermore, higher socioeconomic level was associated with both appropriate antibiotic use for pneumonia and inappropriate for mild ARI. Children in the poorest households in 2011 and 2014 were less likely to appropriately use antibiotics than those from other socioeconomic levels (OR 0.37 and 0.025 respectively). And children in the poorest households in 2014 were less likely to inappropriately use antibiotics for mild ARI than all other socioeconomic levels (OR 0.36).

**Conclusions:** These findings support, challenge, and broaden current understandings of antibiotic usage in Vietnam. Our results suggest that inappropriate antibiotic use arises from the provider and institutional level. Consequently, we argue that community education efforts and enforcing antibiotics as prescription-only is insufficient. Instead, more focus should be made on reducing financial incentives and infrastructural weaknesses at hospitals and health centres. Furthermore, our results show the need to provide the poorest households with sufficient access to antibiotics. Health policy should tackle the issue of inappropriate use of antibiotics for mild ARI among higher socioeconomic groups.

## Background

### Antibiotic Resistance

Antibiotic resistance, the ability of bacteria and other microorganisms to resist the effects of an antibiotic, is a global epidemic (Levy 1998). Antibiotic resistance occurs naturally but exposure to antibiotics accelerates the biological process (Ventola 2015). The prominent forms of misuse in communities and primary health-care in Asia include using antibiotics for viral infections and buying antibiotics over-the-counter without a prescription from a qualified health worker (Levy 1998).

Antibiotic resistance poses a unique, complex, and pressing issue in low- and middle-income countries (LMICs). Since the first introduction of antibiotics, they now constitute the single largest group of drugs purchased in most LMICs (Larsson et al. 2000). The burden of infectious disease remains high in LMICs, accounting for 25% of deaths worldwide and 45% of deaths in LMICs (WHO 2000). Antibiotic resistance is therefore a low priority in LMICs compared to the immediate challenges of infectious diseases (Kotwani and Holloway 2011). The threat of the loss of antibiotics in the future does not pose the same urgency as current deaths. However, without concern for the issue of resistance, current antibiotics will soon be insufficient to combat future epidemics.

Thus, LMICs such as Vietnam face a dual challenge: combatting their high burden of infection and mitigating antibiotic resistance. Research and policy for these nations must consider both the issue of drug access and resistance.

Antibiotic resistance in Vietnam is high. *Escherichia* coli in rural Vietnamese preschool children fecal samples has high resistance to ampicillin (65%), tetracycline (74%) and co-trimoxazole (68%) (Dyar 2012). And in hospital wastewater, 83% of *Escherichia coli* isolates were resistance to at least one antibiotic and 32% were multi- drug resistant (Lien 2017). In Vietnam, 91% of *Streptococcus pneumoniae* isolates were found to be resistant to macrolides and 38% to penicillin (CDDEP 2019). In comparison, in the UK, the resistance percentages for penicillin and macrolides in *Streptococcus pneumoniae* bacteria were 5% and 6% respectively (CDDEP 2019).

### Acute Respiratory Illness in Vietnam

Acute respiratory illness (ARI) prevalence is high in LMICs such as Vietnam. Pneumonia, (ARI with severe symptoms), can be caused by bacteria such as *Streptococcus pneumoniae* and *Haemophilus influenzae*. Without diagnostics such as point-of-care or laboratory testing, antibiotics are prescribed or sold for any respiratory infection with pneumonia-like symptoms. Pneumonia is the 3rd top cause of mortality in Vietnam, the 2nd most common cause of death from infections, and the 2nd leading cause of mortality in children under 5 (GARP, 2010).

### Health Infrastructure

Hospital infrastructure in Vietnam contributes to inappropriate antibiotic use in various ways. One way is due to the rapidly-developing network of over 60,000 private pharmacies and unlicensed drug-sellers, which allow patients to self-medicate, bypassing the public health system to obtain medications. Self-medicating is especially common in rural and lower income individuals, who have limited access to healthcare facilities (GARP 2010). On the other hand, wealthier households have greater access to hospitals, which provide better healthcare and consume a larger share of health services (GARP 2010). Only larger facilities such as provincial and national hospitals have capacity to perform bacterial culture, identification, and susceptibility testing.

Furthermore, there is a shortage of trained staff in the health care sector, with a low nurse-to-doctor ratio, lack of specialists and trained managers, and many vacancies in rural and remote areas. The shortage means that healthcare providers often do not have enough time to educate patients on correct medication usage.

Poor infection prevention and control in hospitals is an additional driver of high antibiotic use, which perpetuates resistance.

### Purchasing Practices

As in many countries, causes of antibiotic misuse in Vietnam include the perceived expectations of patients, physician? time constraints, lack of knowledge, lack of diagnostic capability, and financial benefits for the prescriber (GARP 2010). The availability of antibiotics as over-the-counter drugs and poor enforcement of pharmacy regulations exacerbates misuse. In 2014, after issuing of the 2005 Drug Law, which required antibiotics to be sold with prescription only, antibiotics were sold without a prescription in 88% of randomly surveyed urban drug sale transactions, and 91% of rural transactions (Nga et al. 2014).

Consumer habits may explain why over-the-counter antibiotic sales are so high. Urban and rural respondents report that patients avoid visiting doctors due to inconvenience in cost and time, and prefer visiting private pharmacies for mild diseases (Nga et al. 2014; Duong, Binns, and Le 1997). It is unclear whether this habit applies to children. Any mild illness may cause parental concern and financial constraints are less limited due to universal health insurance for children under 6.

### Economic Incentives

Antibiotics were the most common drug sold in both urban and rural Vietnamese pharmacies (Nga et al. 2014). 31% of urban and 35% of rural sites admitted that the high profitability of antibiotic sales contributed to their inappropriate sale (Nga et al. 2014). And rural and urban pharmacists reported that they feared losing customers and felt pressure from patients to inappropriately dispense antibiotics.

### Knowledge Amongst Healthcare Providers and Caretakers

Misconceptions about antibiotic use has changed in the urban population due to better economic conditions and higher education levels (Nga et al. 2014). However, rural customers consider antibiotics to be a miracle drug and often request it for a private stock for self-medication (Nga et al. 2014). In fact, only 13% of caregivers from Bavi, Vietnam (a rural district of Hanoi) demonstrated correct knowledge of ARI management according to standard guidelines (Hoa et al. 2011). But knowledge was not correlated with caregiver practice (Hoa et al. 2011). Instead, it is likely that caregivers follow healthcare provider recommendations, indicating that healthcare providers play a large role in antibiotic practices (Hoa et al. 2011).

### Self-Medication versus Health Care Provider

It is unclear whether the more important driver of inappropriate antibiotic use is self-medication or health care providers. On one hand, self-medicating individuals may be more likely to use antibiotics than those who seek advice from health professionals (Okumura, Wakai, and Umenai 2002; GARP 2010). On the other hand, seeking healthcare for mild ARI at any health facility may increase the risk of unnecessary antibiotics (Larsson et al. 2005; Hoa et al. 2011). Further investigation using nationwide data is therefore important to clarify the root source of misuse.

### Purpose and Research Questions

No publications that we are aware of have investigated country-wide antibiotic use and care-seeking over time in Vietnam. Nor have there been nationwide investigations for children under 5. Current research has also neglected to assess specific social and healthcare determinants that impact antibiotic use on a country-wide level. Our research aims to fill this gap in knowledge. By further understanding the structural and cultural factors that contribute to antibiotic use in Vietnam, policy- makers can create targeted and effective interventions.

We aim to answer three main questions:

1. Was there a change in antibiotic usage in children under 5 between 2006 and 2014?
2. Was there a change in the type of healthcare provider sought for antibiotics in children under 5 between 2011 and 2014?
3. What are the social and health-seeking determinants of care-seeking, correct antibiotic use and antibiotic misuse?

## Methods

### Data sources

Our research uses publicly available datasets to look at antibiotic usage for ARI in Vietnamese children under 5. We used UNICEF’s Multiple Indicator Cluster Survey (MICS) datasets for the years 2006, 2011, and 2014. These were nationally representative household surveys, conducted using stratified clustered sample designs. Survey data were weighted for all three years in order to better reflect the general population characteristics of Vietnam (UNICEF 2019).

### Indicators and Variable Definitions

We included data for all children under five years for whom data was available. The definitions used for outcome variables, healthcare providers and social determinants are given in detail in Table 1. We attempted to follow the most recent IMCI guidelines as closely as possible and defined self-reported “signs and symptoms of pneumonia” as cough with fast or difficulty breathing, related to a problem in the chest. We were unable to identify signs and symptoms of severe pneumonia as questions about danger signs or stridor were not included in the questionnaire. We were unable to distinguish between bacterial and nonbacterial pneumonia due to lack of confirmatory tests. We defined mild ARI (not pneumonia) as those that either had 1) only cough (and no rapid or difficult breathing), or 2) cough and difficulty breathing but not related to a problem in the chest. Any antibiotic use for mild-ARI cases was classified as “inappropriate antibiotic use.”

**Table 1:** Definitions of Outcome Variables and Social Determinants

| Variable | Definition |
| --- | --- |
| **ARI outcomes** |  |
| Symptoms of pneumonia | Cough with rapid or difficult breathing related to a problem in the chest |
| Mild ARI | Cough with or without difficulty breathing, not related to a problem in the chest |
| **Healthcare Seeking** |  |
| Healthcare provider | Facilities that individuals used for health advice for antibiotics. Included, but not limited to government hospitals, private hospitals, and private pharmacies |
| Government Facility | Government facilities include government health centre, government health post, mobile/outreach clinic, other public |
| Private Facility | Private facilities include private hospital/clinic, private physician |
| Self-medication | Obtaining antibiotics on one's own accord without an additional healthcare provider contact. Includes individuals who visited pharmacies to obtain antibiotics |
| Appropriate antibiotic use | Antibiotic use for symptoms of pneumonia |
| Inappropriate antibiotic use | Antibiotic use for mild ARI |
| **Social determinants** |  |
| Socioeconomic Level | Household wealth quintiles, calculated through principal components analysis of household asset data (grouped into quintiles as lowest, second, third, fourth, highest) |
| Urban/Rural | Region of the child's residence |
| Child Gender | Gender of the child – male or female |
| Child Age | Age of the child, taken in the questionnaire in whole numbers based on date of last birth date |
| Maternal education | Highest level of maternal education achieved, grouped as primary, lower secondary, upper secondary, professional school, college/university. |

### Social Determinants

Healthcare facilities were classified as government facilities, private facilities, and private pharmacies. Individuals who obtained antibiotics on their own accord without going to a formal healthcare provider, are defined as performing “self medication.” This included individuals who went to a pharmacy but no other healthcare provider. Throughout the paper, these individuals will also be distinguished through the term “no resource.” The MICS survey data contained five socioeconomic classes: Lowest, Second, Middle, Fourth, Highest. 2006 MICS Data contain data regarding the gender of the household head. 2006, 2011, and 2014 data all contain gender of the child. We included maternal education as a known determinant of socioeconomic level and child’s health. Education and wealth are correlated but not 100%. The MICS maternal education data contain five categories: Primary, Lower secondary, Upper secondary, Professional school, College/university.

### Statistical Analysis

Logistic regression models were developed to analyze the main effects of social/health-seeking determinants on correct antibiotic use and antibiotic misuse. The exponentiated regression coefficients were interpreted as odds ratios. The independent variables investigated were urban/rural, socioeconomic class, maternal education, child age, child gender, government facility, and private facility. Socioeconomic class and maternal education were treated as categorical variables due to their seemingly non-linear relationship with antibiotic use.

### Ethics

All analyses were based on secondary analysis of publicly available data from national surveys and no separate ethical approval was required.

### Availability of data and materials

The datasets generated and/or analyzed during the current study are available in the UNICEF MICS repository, <u>MICS UNICEF Surveys</u> under Viet Nam.

## Results

### General Characteristics Trends

Children with symptoms of pneumonia decreased from 4.3% of total children under 5 population in 2006 to 1.2% of total children with cough in 2014 (Table 2). And the proportion of children under 5 with symptoms of pneumonia who correctly used antibiotics increased from 63% in 2006 to 83% in 2014 (Table 2). In 2014, the incorrect use of antibiotics for mild ARI was as high as correct use for pneumonia - 82% compared to 83% respectively (Table 2).

**Table 2:** General Characteristics

|  | **2006**<br>n (%) | **2011**<br>n (%) | **2014**<br>n (%) |
| --- | --- | --- | --- |
| **Under 5 Population** | 3,571 | 4,901 | 4,582 |
| **Male** | 1,882 (52.7<sup>^</sup>) | 2,501 (51<sup>^</sup>) | 2,390 (52<sup>^</sup>) |
| **Female** | 1,689 (42.7<sup>^</sup>) | 2,400 (49<sup>^</sup>) | 2,192 (48<sup>^</sup>) |
| **Urban** | 782 (22) | 1,895 (39) | 1,772 (39) |
| **Rural** | 2,789 (78) | 3,006 (61) | 2,810 (61) |
| **Socioeconomic quintile** |  |  |  |
| Lowest | 1,098 (31<sup>*</sup>) | 1,235 (25<sup>*</sup>) | 1,191 (26<sup>*</sup>) |
| Second | 607 (17<sup>*</sup>) | 753 (15<sup>*</sup>) | 783 (17<sup>*</sup>) |
| Middle | 607 (17<sup>*</sup>) | 820 (17<sup>*</sup>) | 839 (18<sup>*</sup>) |
| Fourth | 601 (17<sup>*</sup>) | 985 (20<sup>*</sup>) | 893 (19<sup>*</sup>) |
| Highest | 658 (18<sup>*</sup>) | 1,028 (21<sup>*</sup>) | 1,028 (22<sup>*</sup>) |
| **Maternal Education** |  |  |  |
| None | 556 (16<sup>*</sup>) | 877 (18<sup>*</sup>) | 733 (16<sup>*</sup>) |
| Incomplete primary | 433 (12<sup>*</sup>) | 1,824 (37<sup>*</sup>) | 1,592 (35<sup>*</sup>) |
| Complete primary | 1,138 (32<sup>*</sup>) | 925 (19<sup>*</sup>) | 1,006 (22<sup>*</sup>) |
| Complete lower secondary | 879 (25<sup>*</sup>) | 854 (17<sup>*</sup>) | 897 (20<sup>*</sup>) |
| Complete upper secondary | 565 (15<sup>*</sup>) | 0 | 0 |
| Cough in the past 2 weeks | 962 (27<sup>^</sup>) | 1,842 (38<sup>^</sup>) | 2,095 (46<sup>^</sup>) |
| Symptoms of pneumonia | 155 (4.3<sup>^</sup>) | 75 (1.5<sup>^</sup>) | 54 (1.2<sup>^</sup>) |
| Pneumonia and used antibiotics | 97 (63<sup>**</sup>) | 51 (68<sup>**</sup>) | 45 (83<sup>**</sup>) |
| Mild ARI | 298 (8.3<sup>^</sup>) | 811 (16.5<sup>^</sup>) | 989 (25.6<sup>^</sup>) |
| Mild ARI and used antibiotics | N/A□ | N/A□ | 809 (82<sup>***</sup>) |
| **Source of Care** |  |  |  |
| Government hospital | 49 (5<sup>*</sup>) | 30 (1.6<sup>*</sup>) | 298 (14<sup>*</sup>) |
| Private hospital | 21 (2.1<sup>*</sup>) | 25 (1.4<sup>*</sup>) | 418 (20<sup>*</sup>) |
| Self-medication<br>(includes private pharmacy) | 73 (7.6<sup>*</sup>) | 45 (2.4<sup>*</sup>) | 835 (40<sup>*</sup>) |

<sup>^</sup>Out of under 5 population
<sup>\*</sup>Out of total with cough in past 2 weeks
<sup>\*\*</sup>Out of total with pneumonia
<sup>\*\*\*</sup> Out of total with mild ARI
□ Not included in survey

### Determinants of Appropriate Antibiotic Use (for cough with pneumonia-like symptoms)

Children from households in the highest socioeconomic level in 2006 and 2011 were about 4 times more likely than all other levels to use antibiotics for pneumonia (Table 3). In 2011, children in the fourth socioeconomic level (second highest) were 7 times more likely to use antibiotics for pneumonia. On the other hand, children from the lowest and second-lowest socioeconomic levels in 2011 were 63% and 73% less likely to use antibiotics for pneumonia (Table 3). In 2014, the poorest families were 98% less likely to use antibiotics for pneumonia (Table 3).

**Table 3:**
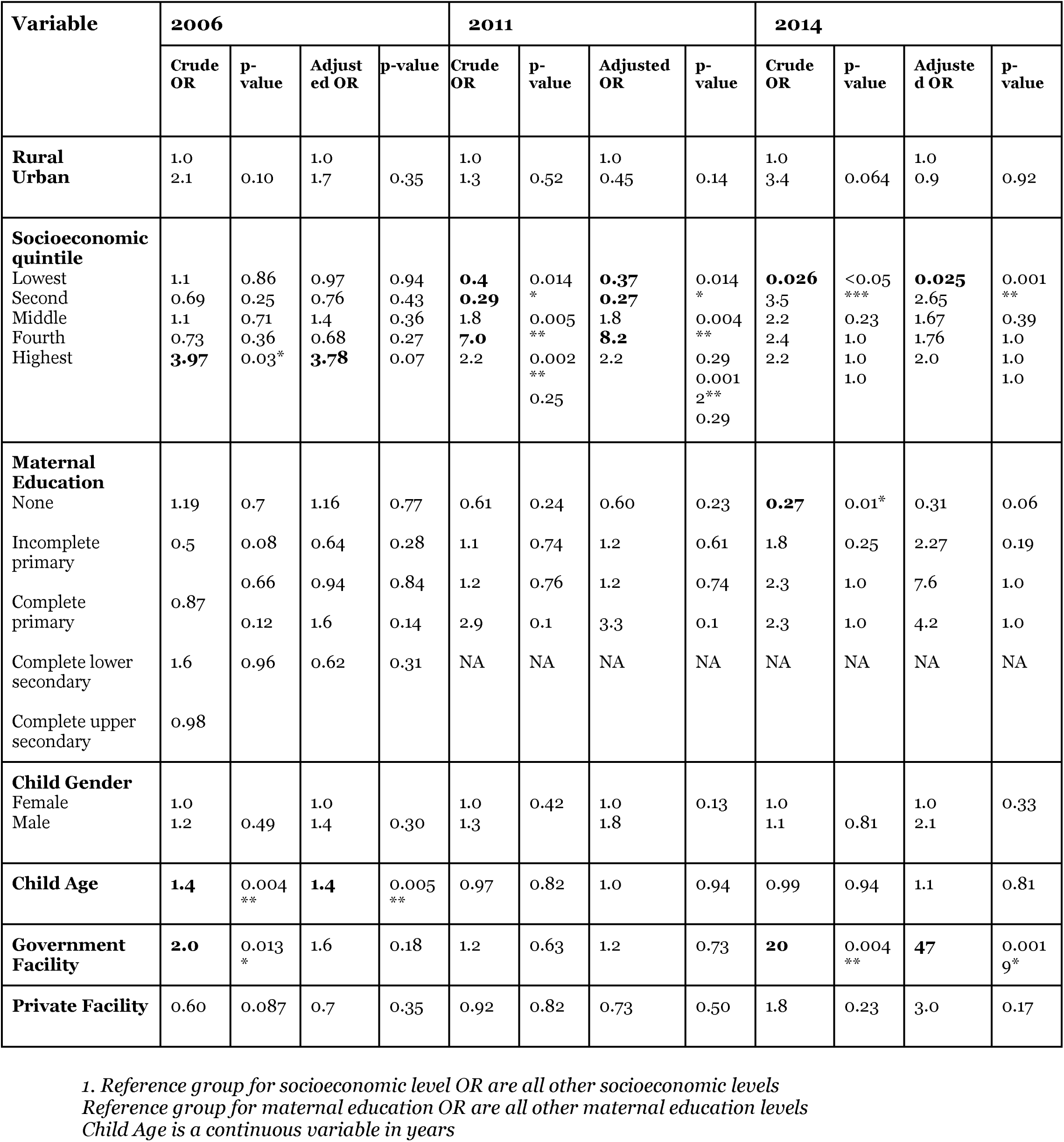
Determinants of Appropriate Antibiotic Use Among Children with Symptoms of Pneumonia (in Odds Ratio)

A higher proportion of children from households with a female head used antibiotics for pneumonia than those in households with a male head in 2006 (83% versus 54% respectively) (Figure 1).

**Figure 1:**
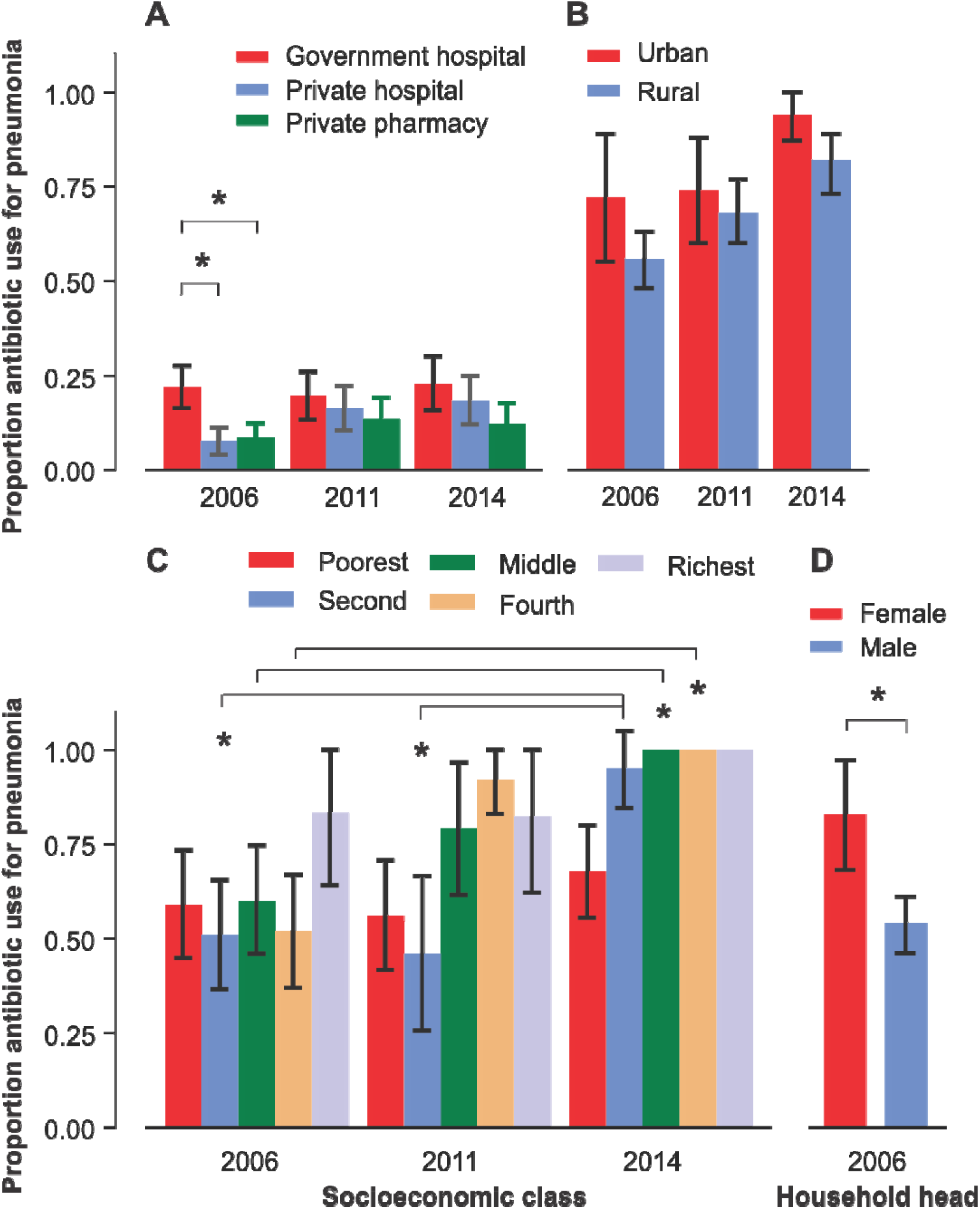
Appropriate Antibiotic Use for Pneumonia in Children Under 5. <u>A) Appropriate antibiotic use over time for different healthcare provider types: government hospital, private hospital, and private pharmacy.</u> *Proportion calculated from the number of children who used antibiotics for pneumonia in particular facility out of total children with pneumonia <u>B) Appropriate antibiotic use over time for urban / rural regions</u> *Proportion calculated from the number of children who used antibiotics for pneumonia in rural or urban region out of total children with pneumonia in urban or rural region <u>C) Appropriate antibiotic use over time for different socioeconomic levels (out of those with pneumonia in the socioeconomic level)</u> *Proportion calculated from the number of children who used antibiotics for pneumonia in each socioeconomic level out of total children with pneumonia in each socioeconomic level <u>D) Antibiotic use in female versus male household heads for 2006.</u> *Proportion calculated from the number of children who used antibiotics for pneumonia in either female or male household head out of total children with pneumonia in either female or male household head

### Determinants of Inappropriate Antibiotic Use (for mild ARI)

Children who sought care from a health care provider had a significantly higher proportion of antibiotic use for mild ARI than those who did not (86% versus 71%) (Figure 2). The higher misuse in those who sought care is present in both urban and rural children: 81% versus 49% in urban settings, and 83% versus 36% in rural settings (Figure 2).

**Figure 2:**
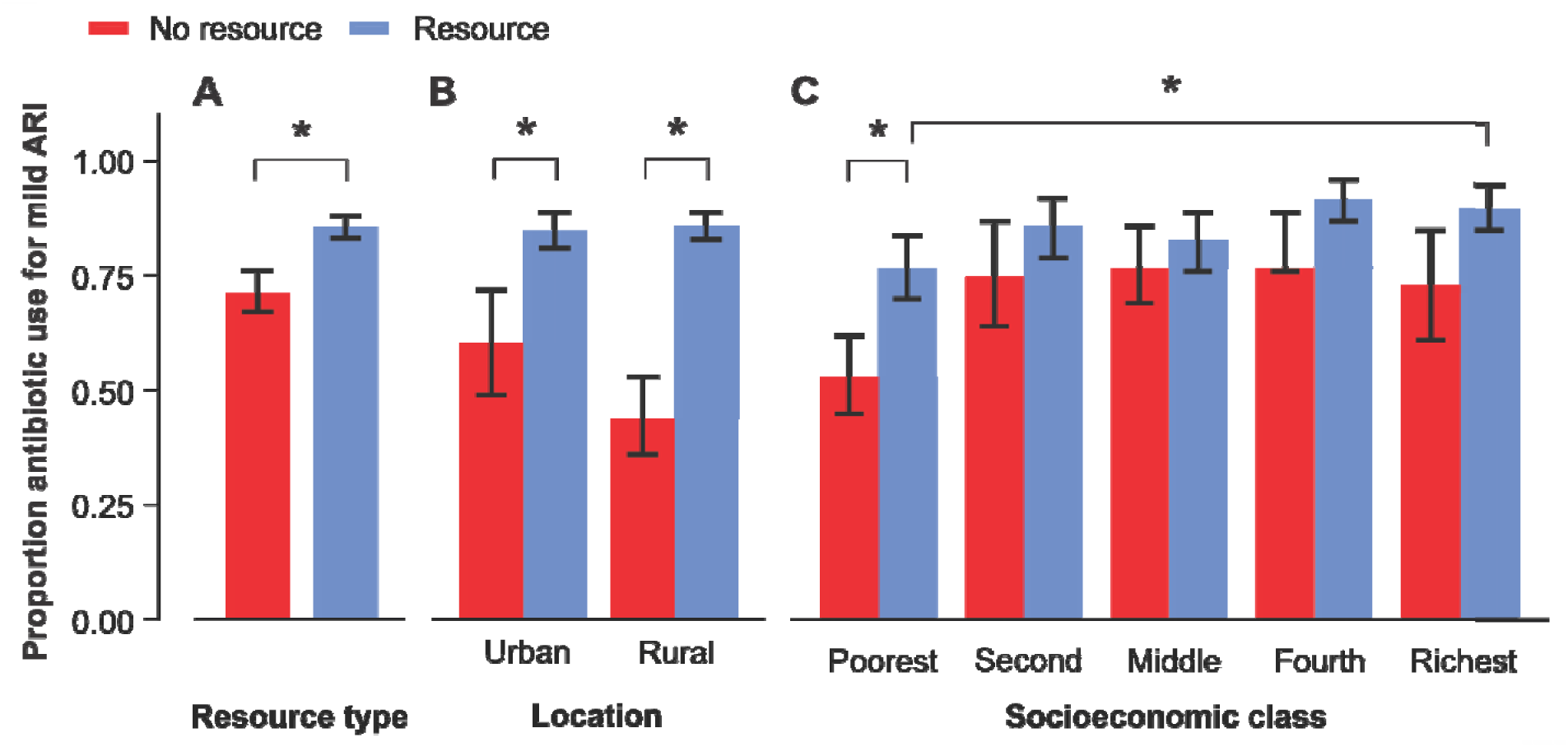
Inappropriate Antibiotic Use for Mild ARI in 2014. <u>A) General inappropriate antibiotic use for mild ARI proportions in no resource versus resource individuals.</u> *For no resource category, proportion was calculated from the *number of children who used antibiotics for mild ARI with no resource / children with mild ARI with no resource*. For resource category, proportion was calculated from *number of children who used antibiotics for mild ARI with resource / children with mild ARI with resource*. <u>B) Inappropriate antibiotic use for mild ARI in urban and rural regions, for individuals with no resource versus resource</u> *For the urban category, proportion was calculated from the *number of children in urban regions who used antibiotics for mild ARI with no resource / children with mild ARI in urban regions with no resource*. For the rural category, proportion was calculated from *number of children who used antibiotics for mild ARI with resource / children with mild ARI with resource*. <u>C) Inappropriate antibiotic use for mild ARI in socioeconomic levels, for individuals with no resource versus resource</u> *For no resource category, proportion was calculated from the *number of children who used antibiotics for mild ARI with no resource / total children with mild ARI with no resource. For resource category, proportion was calculated from number of children who used antibiotics for mild ARI with resource / children with mild ARI with resource*.

The highest socioeconomic level was more likely to use antibiotics for mild ARI (OR = 1.6, p = 0.07) (Table 4). And the lowest socioeconomic level was less likely than all other socioeconomic classes to use antibiotics for mild ARI (OR = 0.36, p < 0.005) (Table 4). Children who sought care at private facilities were 6 times more likely than all other socioeconomic classes to misuse antibiotics (OR = 6.1, p < 0.005) (Table 4). Children who sought care at government facilities were 3.8 times more likely to misuse (OR = 3.8, p < 0.005) (Table 4).

**Table 4:** Determinants of Inappropriate Antibiotic Use for Mild ARI

| Variable | 2014 |  |  |  |
| --- | --- | --- | --- | --- |
|  | Crude OR<br>(95% CI) | p-value | Adjusted OR<br>(95% CI) | p-value |
| **Rural** | 1.0 |  | 1.0 |  |
| **Urban** | 1.26 | 0.15 | 0.75 | 0.15 |
| **Socioeconomic** |  |  |  |  |
| Poorest | **0.32** | $< 0.005^{***}$ | **0.36** | $< 0.005^{**}$ |
| Second | 2.3 | 0.27 | 1.3 | 0.32 |
| Middle | 0.95 | 0.79 | 0.88 | 0.55 |
| Fourth | 2.8 | $0.005^{**}$ | 2.6 | $< 0.005^{***}$ |
| Richest | 1.7 | $0.02^*$ | **1.6** | 0.07 |
| **Maternal Education** |  |  |  |  |
| None | 0.92 | 0.7 | 0.88 | 0.59 |
| Incomplete primary | 1.0 | 0.78 | 1.0 | 0.94 |
| Complete primary | **1.5** | $0.04^*$ | 1.4 | 0.12 |
| Complete lower secondary | 1.1 | 0.62 | 1.14 | 0.56 |
| Complete upper secondary | NA |  | NA |  |
| **Child Gender** | 0.97 | 0.84 | 1.0 | 0.86 |
| **Child Age** | **1.2** | < 0.005** | **1.2** | 0.01* |
| **Government Facility** | 2.4 | 0.13 | **3.8** | < 0.005*** |
| **Private Facility** | **4.2** | < 0.005*** | **6.1** | < 0.005*** |

1. \*Adjusted Odds Ratio (OR) regressions include the following variables: socioeconomic, child age, child gender, urban, maternal education, government facility, private facility 2. Signif. codes: 0 '\*\*\*' 0.001 '\*\*' 0.01 '\*' 0.05 '.' 0.1 ' ' 1 3. Socioeconomic class as numerical: poorest = 1, second = 2, middle = 3, fourth = 4, richest = 5 4. Maternal education as numerical: none = 1, Incomplete primary = 2, Complete primary = 3, complete lower secondary = 4, complete upper secondary = 5

## Discussion

Our nation-wide analysis indicates that Vietnamese healthcare providers may be perpetuating, rather than mitigating, antibiotic misuse. Thus, inappropriate antibiotic use in Vietnam is not only due to lack of knowledge in the community but also provider and institutional level problems. Our results also show disparities in antibiotic use amongst socioeconomic levels, with the lowest socioeconomic levels still less likely to obtain antibiotics for pneumonia than higher levels. However, higher socioeconomic levels were not only more likely to correctly use antibiotics but also to overuse the drugs for mild ARI cases.

### Healthcare Provider

Our results suggest that those who seek advice from a healthcare provider are more likely to use antibiotics for mild ARI (Figure 2, Table 2). This is evident in the lowest socioeconomic level, and in both rural and urban settings (Figure 2). The results complicate the current assumptions that self-medication in low-income rural areas is a contributing factor to inappropriate antibiotic use (GARP 2010). Our nation-wide analysis indicates that Vietnamese healthcare providers may be contributing to antibiotic misuse rather than reducing it. So misuse is due to not only lack of knowledge in the community but also issues such as financial incentives in providers and infrastructural inadequacies.

### Socioeconomic Level

Lower proportions of children from low socioeconomic households are using antibiotics for pneumonia compared to higher socioeconomic levels. The pattern of increased use between 2006 and 2014 reflected “elite capture”, with the most well-off families gaining better access first. In 2014, among all socioeconomic levels except the poorest, more than 80% of children with pneumonia received antibiotics (Figure 1C). Antibiotic access increased first for the highest socioeconomic level, with levels already high in 2006. The second highest socioeconomic level experienced a large increase in antibiotic usage for pneumonia from 2006 to 2011 (51% to 95%). The middle socioeconomic level increased their antibiotic use more gradually between 2006 and 2011 and again between 2011 and 2014. The second-lowest socioeconomic level had their largest increase in antibiotic usage later, from 2011 to 2014 (46% to 95%). However, the lowest class did not change in their correct antibiotic usage between 2006 and 2014, with less than 75% of children obtaining antibiotics. Logistic regression results support this descriptive analysis, with the highest socioeconomic level more likely to use antibiotics for pneumonia in 2006 and 2011 compared to all other levels (Table 2). And children from the lowest and second-lowest levels are less likely to correctly use antibiotics compared to those of other levels. Thus, in the fight against antibiotic overuse and resistance, it is important to remember that there are children from poor families who still fail to obtain antibiotics when needed.

Antibiotic use for mild ARI (inappropriate use) is also significantly higher in the highest socioeconomic level compared to the lowest (Figure 2C). And children from the highest socioeconomic levels are 1.6 times more likely to inappropriately use antibiotics compared to other socioeconomic levels, while children from the lowest socioeconomic levels are 64% less likely to misuse antibiotics compared to other levels (Table 4). These results indicate that having the financial means to obtain antibiotics not only increases propensity to use antibiotics for pneumonia (correct use) but also to use them for mild ARI (inappropriate use). This trend may reflect the cultural view of antibiotics as a cure-all. It also suggests a lack of knowledge of correct antibiotic usage in higher socioeconomic levels in spite of their higher education levels.

### Household Head

A higher proportion of children in households with a female head used antibiotics for pneumonia than those in households with a male head (83% versus 54% respectively). It is possible that households with a female head have other characteristics that make it more likely for their child to have access to antibiotics. Further research is needed to clarify this result.

### Urban-Rural Divide

No significant differences in correct antibiotic use and misuse were found in urban versus rural locations. The lack of significant differences suggest that antibiotic use for children is more impacted by socioeconomic constraints than barriers in transportation and physical access.

### Broader Implications

Overall, this paper’s nationwide results show that healthcare-seeking and high socioeconomic levels increase the propensity for inappropriate antibiotic use. Therefore, improving antibiotic stewardship efforts in health departments, hospitals, and healthcare providers are just as important as improving community-level understanding. Of course, it is impossible to completely disentangle healthcare inadequacies from insufficient information at the community level. A trickle-down effect is likely to occur, wherein infrastructural inadequacies contribute to misinformation in the community. Patient expectations are also known to contribute to incorrect prescription practices (Vinker, Ron, and Kitai 2003; Macfarlane et al. 1997; Mangione-Smith et al. 2001). Furthermore, parenting style may contribute to a distinct parental demographic in those who sought care from a provider. This nuance is a potential confounding influence, but does not detract from the reality of the general trend.

### Limitations

There were some limitations to this analysis. Although UNICEF surveys use similar methodology and questionnaires at each time point, there were some small differences. Only 2014 data provided information about inappropriate antibiotic use for mild ARIs. In 2006 and 2011 questionnaires, the question about antibiotic usage was skipped for those with mild ARI.

Different seasons will affect disease prevalence especially in ARI, which are greater during the wintertime. Only 2014 surveys indicated the season (Dec 2013 – April 2014) during which the questionnaires were distributed. Therefore, our comparison of 2006, 2011, and 2014 general trend characteristics were made under the assumption that the 2006 and 2011 surveys were conducted during similar seasons. However, given that there is a decreasing trend in ARI and the 2014 survey was conducted during the peak ARI season, decreases in ARI over time are likely to be real. Furthermore, antibiotic use decisions for those with ARI should not be influenced by season.

Furthermore, the UNICEF data provides information only on whether a child obtained antibiotics for pneumonia-like symptoms or mild ARI. The duration of antibiotic usage is unknown. Therefore, “correct” antibiotic use assumes that anyone who obtained antibiotics for ARI is using it according to prescription instructions. This paper could not assess lack of adherence to drug instructions as another form of antibiotic misuse. The data also lacks the class and type of antibiotic use, so we are unable to discern whether the narrow or broad spectrum antibiotics were used.

Additional limitations of the research stem from the nature of survey data. Survey data is self-reported to the interviewer, so response bias and inaccurate caretaker recall are possible. Furthermore, pneumonia and ARI are diagnoses normally made by medical professionals that may not fully correlate with the indicators used here.

## Conclusion

Our findings support, challenge, and broaden current understandings of antibiotic usage in Vietnam. In fact, our results suggest that inappropriate antibiotic use arises from the provider and institutional level. Consequently, we argue that community education efforts and enforcing antibiotics as prescription-only is insufficient. Instead, more focus should be made on reducing financial incentives and infrastructural weaknesses at hospitals.

Furthermore, even in the midst of antibiotic overuse and resistance, the issue of antibiotic access should not be overlooked. Policy efforts should improve access to primary care, such as transportation and hours of service, in order to increase access to antibiotics for those who need it in low income groups.

## Data Availability

The datasets generated and/or analyzed during the current study are available in the UNICEF MICS repository, MICS UNICEF Surveys under Viet Nam.

http://mics.unicef.org/surveys

## Acknowledgements

A big thank you to the Oxford Clinical Research Unit team in Hanoi, Vietnam for their friendship and support.

## List of abbreviations

ARI: Acute Respiratory Infection
MICS: Multiple Indicator Cluster Survey
UNICEF: United Nations Children’s Fund
WHO: World Health Organization
LMICs: Low- and Middle-Income countries

